# Quantifying transmissibility of COVID-19 and impact of intervention within long-term health care facilities

**DOI:** 10.1101/2021.02.01.21249903

**Authors:** Jessica E. Stockdale, Sean C. Anderson, Andrew M. Edwards, Sarafa A. Iyaniwura, Nicola Mulberry, Michael C. Otterstatter, Naveed Z. Janjua, Daniel Coombs, Caroline Colijn, Michael A Irvine

## Abstract

Estimates of the basic reproduction number (*R*_0_) for Coronavirus disease 2019 (COVID-19) are particularly variable in the context of transmission within locations such as long-term health care (LTHC) facilities. We sought to characterise the heterogeneity of *R*_0_ across known outbreaks within these facilities. We used a unique comprehensive dataset of all outbreaks that have occurred within LTHC facilities in British Columbia, Canada. We estimated *R*_0_ with a Bayesian hierarchical dynamic model of susceptible, exposed, infected, and recovered individuals, that incorporates heterogeneity of *R*_0_ between facilities. We further compared these estimates to those obtained with standard methods that utilize the exponential growth rate and maximum likelihood. The total size of an outbreak varied dramatically, with a range of attack rates of 2%–86%. The Bayesian analysis provides more constrained overall estimates of *R*_0_ = 2.19 (90% CrI [credible interval] 0.19–6.69) than standard methods, with a range within facilities of 0.48–10.08. We further estimated that intervention led to 57% (47%–66%) of all cases being averted within the LTHC facilities, or 73% (63%–78%) when using a model with multi-level intervention effect. Understanding the risks and impact of intervention are essential in planning during the ongoing global pandemic, particularly in high-risk environments such as LTHC facilities.

## 1. Introduction

Early outbreaks of coronavirus disease 2019 (COVID-19) occurred in locations including public transit, places of worship, cruise ships, meat-packing plants, ski resorts, prisons, and fishing vessels [1, 2, 3, 4, 5, 6, 7]. Prisons have also been highlighted as potential sources of high transmission due to housing an overcrowded aging population with underlying health conditions [8]. Similarly, detention facilities have been noted as locations where rapid transmission could lead to hospitalizations exceeding local healthcare capacity [9]. Long-term health care (LTHC) facilities have also been sites of large COVID-19 outbreaks [10, 11]. The virus has been observed to spread rapidly in the clustered susceptible population of LTHC facilities. Since most people in LTHC are old and frail, morbidity and mortality during an outbreak is often very high: with case fatality rates around 30% [12, 13].

There has been considerable focus on estimation of the basic reproduction number *R*_0_ in different jurisdictions during the COVID-19 pandemic [3, 14, 15, 16, 17, 18]. Given the high fatality rate, there is a keen interest in assessing the impact of COVID-19 on LTHC facilities [19, 20, 13, 21, 22]. However, to date there has been limited work on estimation of reproductive numbers or other transmission parameters within LTHC or other high-risk facilities. Estimates of transmission parameters are useful for retrospective analysis of the efficacy of interventions — particularly when preventative measures have changed between facilities or over time within one facility — and also in planning for future outbreaks. By comparing facility-specific estimates to those from the general population, we can also understand how transmission in LTHC environments differs.

Within British Columbia (BC), Canada, there were 99 identified and defined ‘reportable’ outbreaks, including LTHC facilities, other acute care or assisted living facilities, workplaces, correctional facilities, and religious institutions by the end of September 21 2020. Fifty-three of these were in LTHC facilities. Protocols for LTHC outbreak management have been established, including implementation of a suite of interventions once an outbreak has been identified within a facility [23, 24]. These interventions include isolation of active cases, isolation of all residents in the facility, increased measures around infection control, reduced barriers to testing and increased testing frequency, and increased staffing and resources. Although the precise timing of the implementation of these interventions can vary, they typically occur one to two days after identification of a resident or staff case of COVID-19.

In this work, we estimate *R*_0_ in British Columbia LTHC COVID-19 outbreaks using a dynamic susceptible-exposed-infected-recovered (SEIR) model within a Bayesian hierarchical framework. A time-dependent infection rate in the SEIR model incorporates the implementation of COVID-19 interventions upon identification of each outbreak. The hierarchical framework allows for each LTHC outbreak to be analysed within the context of all other outbreaks in the dataset, and is therefore useful for small outbreaks where the information available from each is limited. Use of a Bayesian model also allows for the incorporation of prior knowledge into the analysis, as well as the ability to fully explore the posterior distribution of the estimates. We compare the estimates from the Bayesian hierarchical model to those from several established statistical approaches for *R*_0_ estimation, which consider each LTHC outbreak independently.

## 2. Methods

### 2.1. Data

Data on reported outbreaks within BC were identified through the BC Centre for Disease Control. Those outbreaks that were identified as having taken place within a LTHC facility and with more than one case were selected. The data consist of reported date, symptom onset date, and facility location of all identified cases of COVID-19.

Dates of infection onset were constructed from reported symptom onset date. Missing symptom onset dates (44/536 cases across all facilities) were replaced by selecting a symptom onset date uniformly at random from all known symptom onset dates within the same facility. Sensitivity analysis to this interpolation was performed, and results are included in the Supplemental Materials (Table S1). The start time of each outbreak was selected as the earliest symptom onset date within all identified cases in that outbreak. The reported date for each outbreak was recorded as the earliest reported date among all reported cases in the facility, since in BC one case in a LTHC facility is sufficient to be labelled as an outbreak.

Additional covariate data concerning the LTHC facilities included in this study was obtained from the Office of the Seniors Advocate, British Columbia website [25]. Where available, we compiled data on various factors for each facility, including average resident age, average resident stay and year of facility opening. These data are from 2018/19: the most recent year available.

### 2.2. Bayesian hierarchical model

We introduce a novel modelling framework for estimating *R*_0_ in LTHC facilities, by incorporating a dynamic compartmental SEIR model into a Bayesian hierarchical structure that includes both parameter and observation uncertainty. This framework incorporates data on each outbreak, allowing for a more robust approach to estimation than considering each outbreak separately, such that outbreaks with few cases can depend strongly on a prior distribution of *R*_0_.

Previous modelling work has, for example, incorporated a chain binomial transmission model within a Bayesian hierarchical model in order to incorporate multiple households into one framework [26], or sought to estimate *R*_0_ within a Bayesian hierarchical framework using the final size of an outbreak rather than incorporating a disease dynamic model [27]. Disease dynamic models have also been used within Bayesian hierarchical frameworks for forecasting seasonal influenza [28]. Our methodology combines the flexibility of dynamic compartmental models for transmission, allowing for estimation not just of the underlying transmission parameters but also the strength of interventions, with the benefits of Bayesian hierarchical fitting such as increased robustness of estimates and incorporation of prior knowledge.

The baseline transmissibility of COVID-19 in our model is assumed to vary between facilities due to differences in layout, contact structure, and demographics of the facility. Within BC, standard policies seeking to reduce transmission were implemented across the province. We therefore model the interventions as having equal strength in all facilities under the assumption that implementation occurs within one day of the first reported case. We also perform a sensitivity analysis in which the intervention strength is allowed to vary between facilities.

#### 2.2.1. Transmission model

We begin with a standard SEIR model where the transmission term in a facility is dependent on the time since identification of the first case in the facility. An individual in facility *k* starts out as susceptible to COVID-19 (*S*_*k*_), following an infection they transition to an exposed group (*E*_*k*_), after an incubation period the individual transitions to an infected group (*I*_*k*_), before finally transitioning to a recovered group; *S*_*k*_, *E*_*k*_, and *I*_*k*_ represent the number of individuals in each group. Due to the short nature of a facility outbreak, imported and exported cases are not considered. Each outbreak is considered to be initiated by a single infectious individual. During the period of this study, general transmission in the community was low, giving a small risk of further importation over a short period. We incorporate uncertainty into the facility population sizes in part to account for staff, but otherwise do not model staff-resident contact any differently than resident-resident contact.

The model equations can be represented as

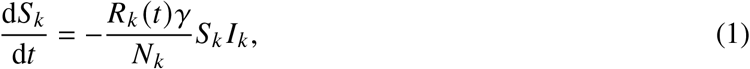

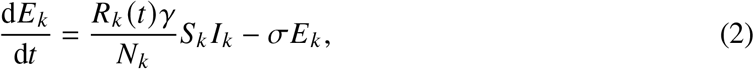

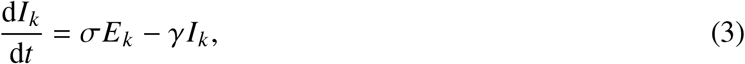

where *R*_*k*_ (*t*) is the reproduction number in facility *k* at time *t, γ* is the recovery rate, *σ* is the incubation period and *N*_*k*_ is the population size, which is approximated with the capacity of facility *k*. After detection of the first case, interventions are implemented aiming to reduce the infectiousness within an outbreak cluster. We model this with a modified *R*_*k*_ (*t*) term that incorporates the initial reproduction number *R*_0,*k*_ in facility *k*, and the modified reproduction given intervention. We assume that an outbreak will eventually be brought under control leading to *R*_*k*_ (*t*) < 1. We therefore define *R*_*k*_ (*t*) using the form

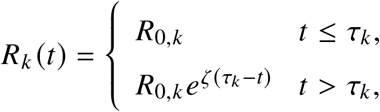

where *ζ* is the intervention effect rate and *τ*_*k*_ is the time of the first case reported within the facility.

#### 2.2.2. Parameter model

The parameter model incorporates the above transmission model with a multi-level *R*_0_ term that is informed by data from all LTHC facilities in the study. More concretely, for outbreak *k*, an *R*_0,*k*_ is drawn from the distribution:

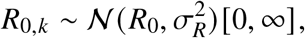

where *R*_0_ is the population-level basic reproduction number and the standard deviation *σ*_*R*_ is drawn from a standard half-normal prior [29].

Literature-based priors were selected for the population-based reproduction number *R*_0_, the infectious period (recovery rate), and the incubation period. As these parameters represented an estimate of the population mean, their variances were characterised by the literature-based standard error making them highly informative. The mean rate of incubation prior (*σ*) had a mean of 0.2 (5 days) and standard deviation 0.025 [30]. The mean rate of recovery (*γ*) had a prior mean of 0.125 (8 days) with a standard deviation of 0.0125 (providing a 95% CI of approximately 6.7–10.0 days). The prior for *R*_0_ was given a mean of 3.0 with a standard deviation of 1.0 to account for a population mean that may be higher than observed in general community transmission [31]. Other parameters, including the between-facility variance in *R*_0_, 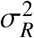, and timing and strength of the intervention, had more uncertainty in their prior distributions. To capture uncertainty in the facility population sizes, the population size of facility *k, N*_*k*_, was given a normally distributed prior with mean equal to the known facility capacity 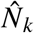 and standard deviation 10. The prior for the initial proportion of susceptible individuals *S*_*k*_ (0) was selected such that the mean would be one individual in a population of 100 with a standard deviation of one person.

The full Bayesian model (without the data likelihood, which is described below) is as follows. All rates have units days^−1^.

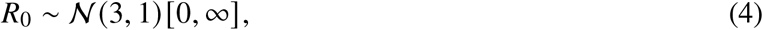

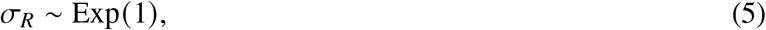

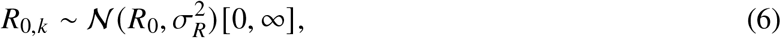

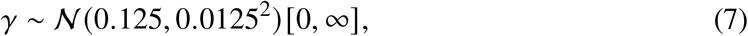

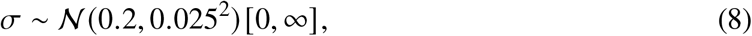

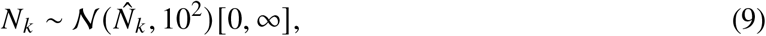

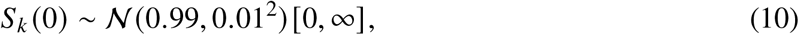

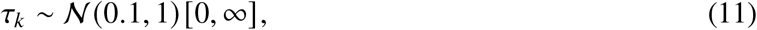

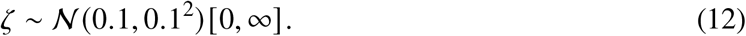

The likelihood is constructed by treating the modelled daily incidence for each facility as a Poisson-distributed random variable, with the rate given as the rate of transition from the *E*_*k*_ to *I*_*k*_ class (ι_*k*_ (*t*)). Data on reported onset of symptoms on a given day for each facility (*C*_*k*_ (*t*)) was used as opposed to date of test, as often tests were conducted en masse at a facility once a primary case had been identified. The cases were then sampled according to the following to construct the likelihood,

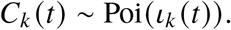

Sampling of the posterior was performed with No U-Turn Sampling (NUTS) [32] using four chains, a warm-up period of 300 samples, and 600 iterations. Visual inspection of the chains, pair-plots, Gelman-Rubin (split-chain-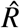) statistic, and presence of divergent transitions were used to assess convergence and mixing [33]. All Bayesian analysis was conducted using the Stan library [34], and our model is provided as an R package accompanying this article (see data access section).

As a sensitivity analysis to understand the potential heterogeneity of the effect of intervention, the model was extended to include a hierarchical structure for the intervention effect *ζ*. The hyper-priors were kept the same as the *ζ* prior in the original model, with inter-facility variance *σ*_*ζ*_ exponentially distributed with rate 1.

Counterfactual scenarios were performed on all facilities by drawing a set of parameters from the posterior and then setting the intervention effect *ζ* to zero for the counterfactual parameter set. We numerically solved the SEIR model for each parameter set and sampled from the model to produce a difference in total incidence between the two scenarios.

Validation of the hierarchical Bayesian procedure was also performed using simulated data. We simulated 30 outbreaks from an SEIR model, each with *R*_0,*k*_ drawn from a 𝒩 (3, 9) distribution truncated at 0, Poisson drawn incidence, and a fixed intervention implementation. Each *R* _0,*k*_ was then estimated within the Bayesian hierarchical framework described above.

### 2.3. Other R_0_ estimation methods

Established methods for estimating *R*_0_ include calculation from the attack rate, which is the total proportion of a given fixed population that contracted the disease during an outbreak [35]. Here, we estimated the attack rate as the total number of reported cases in the outbreak divided by the maximum capacity of the facility. The relationship between attack rate and *R*_0_, under the model of the general deterministic epidemic, is given through the transcendental equation [36, 37]

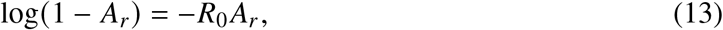

where *A*_*r*_ is the attack rate. We estimated *R*_0,*k*_ for each facility from the attack rate using Equation 13 with *A*_*r*_ = *A*_*r,k*_ and *R*_0_ = *R*_0,*k*_. As a sensitivity analysis the number of exposed individuals (the denominator in the attack rate) was varied between 85% and 115% of the maximum capacity for each facility. However, this approach does not take into account the fact that interventions were put in place during the outbreak, and so resulting estimates are not true basic reproduction numbers but instead represent an averaging across the entire outbreak.

The basic reproduction number in each outbreak was estimated using two other approaches which consider the early growth period of an outbreak only: from the exponential growth (EG) rate [38] and from maximum likelihood (ML) [39]. These methods require an assumed generation time, for which a Gamma distribution with mean 5.2 and standard deviation 1.73, as estimated for COVID-19 in [40], was used. The EG method calculates the exponential growth rate *r*_*k*_ from the initial exponential period of outbreak *k*, which we define as the time from symptom onset of the first case to the maximum incidence day. Poisson regression is used to estimate *r*_*k*_, to account for the fact that the incidence data are integer valued. *R*_0,*k*_ is then calculated as 1/*M* (−*r*_*k*_), where *M* is the moment generating function of the generation time distribution. The ML method assumes that the offspring distribution is Poisson distributed with expectation *R*_0,*k*_. *R*_0,*k*_ is estimated by maximising the resulting Poisson likelihood, given daily incident case counts. This estimation is also performed over an initial exponential period only, which we define as in the exponential growth method. Both the exponential growth and maximum likelihood approaches were implemented using the R0 library in R [41].

All analyses were run using R version 4.0.0 [42, 43].

## 3. Results

Data on 99 outbreaks within BC, Canada, as of September 21 2020 were identified. Of these identified outbreaks, 53 were selected as having taken place within a LTHC facility. Of these, 4 were excluded as they had ambiguous names or were otherwise not able to be linked to the facility characteristics data, 30 were excluded as only having one case, and 1 was excluded as still ongoing, resulting in 18 identified LTHC outbreaks.

Outbreak duration and size varied greatly between locations with a duration of outbreak between 10–55 days (Fig. 1) and size of outbreak between 3 and 89 cases (Fig. 2 & Table S2), leading to an overall attack rate between 2–86% (Table S2). Outbreak size variation showed no clear pattern by reported facility size (Fig. 2).

**Figure 1:**
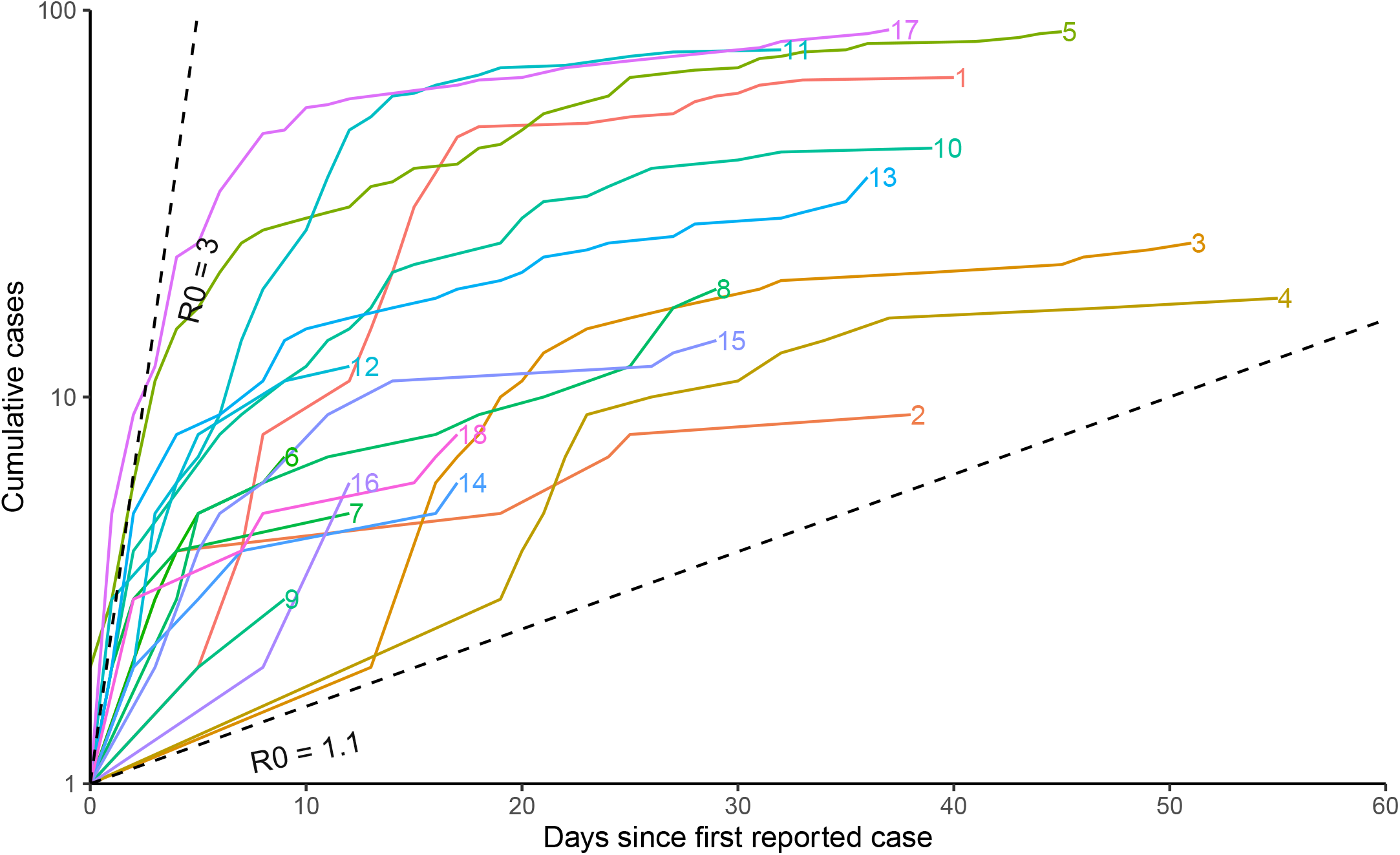
Cumulative cases by LTHC outbreak in BC, Canada. Cumulative number of cases for each identified outbreak by symptom onset, labelled with the location number. Early growth of cases will follow a straight line on a semi-log scale if growth is exponential. Cumulative cases are displayed as linearly increasing between successive symptom onset dates within the facility, rather than with discrete jumps. Dashed lines represent unimpeded epidemics with *R*_0_ values of 1.1 and 3, with a serial interval of 5 days.

**Figure 2:**
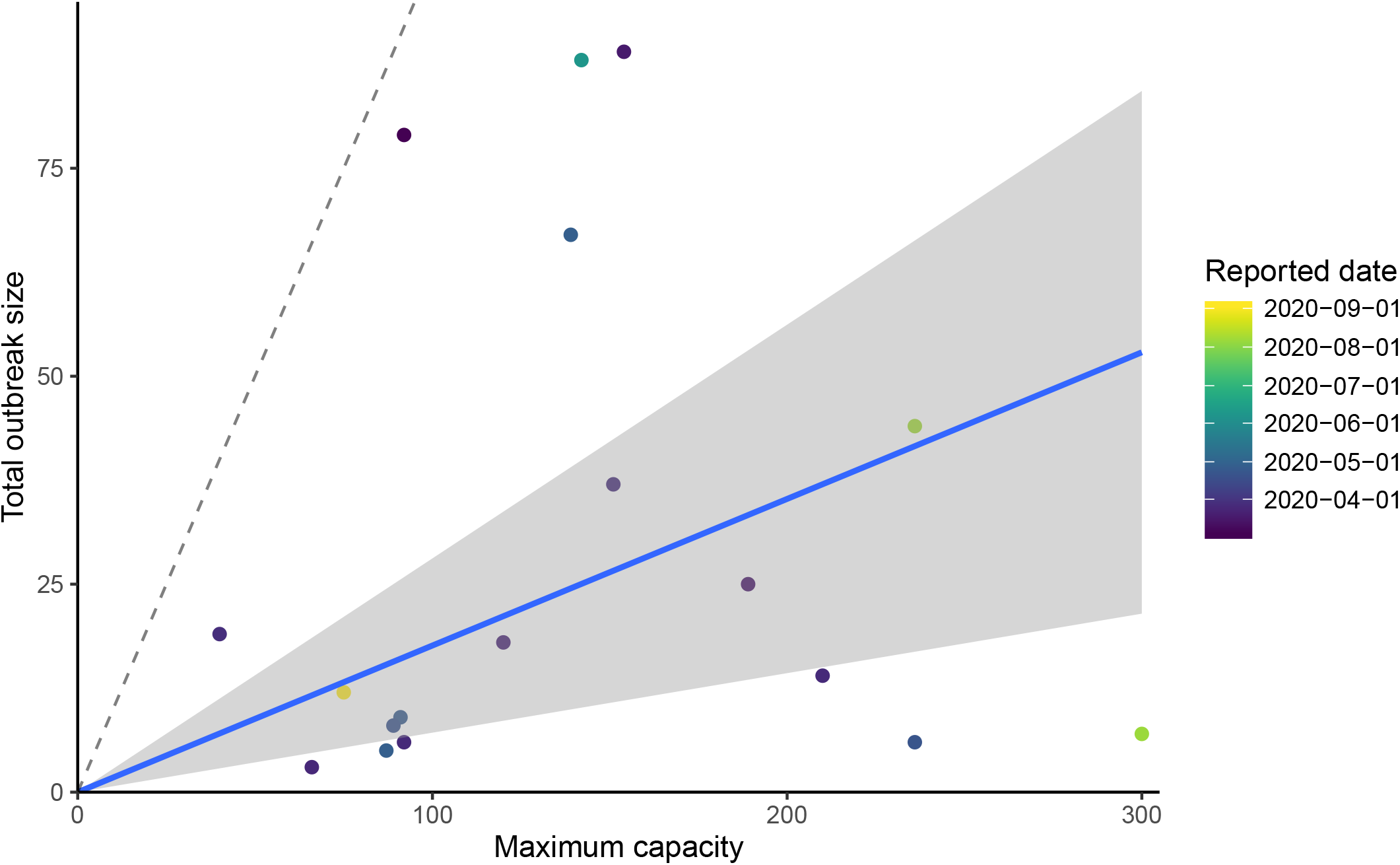
Total outbreak size by facility capacity of LTHC facilities in BC, Canada. Total outbreak size of LTHC facilities are shown by the reported maximum capacity of that facility. A linear regression with zero intercept was fit to these data to produce an estimated overall attack rate of 11.0% (95% CI 0.04%–17.6%), shown by the blue line and shaded areas. Dashed line is the one-to-one line.

Bayesian hierarchical model fitting of *R*_0_ found a large range of estimates by location, with 4 of 18 locations having median *R*_0,*k*_ below the critical threshold of 1, but all of these having 1 within the confidence interval (Fig. 3, Fig. 4 & Table S2). The overall predictive *R*_0_ mean estimate is 2.19 (90% CrI 0.19–6.69), however, the range of *R*_0,*k*_ estimates by location varied beyond this interval between the lowest (location 6) 0.48 (0.08–2.36) to the highest (location 11) 10.08 (7.83–13.02). The incorporation of a hierarchical structure for the intervention strength (*ζ*) increases the overall estimate of *R*_0_: the mean estimate becomes 2.86 (0.35–7.97) (Fig. S1, Fig. S2 & Table S2). We also estimated the ‘critical time’ in each facility, that is, we sampled from the posterior the length of time that it took for the reproduction number to fall from *R*_0,*k*_ to below 1 once each outbreak was reported and full-scale interventions were implemented. The critical time varies with *R*_0,*k*_, but ranged between 0.0 (for outbreaks with *R*_0,*k*_ already below 1) and 4.96 days (Fig. 5 & Table S2). Counterfactual modelling was conducted to estimate the impact of intervention on the total number of cases averted compared to an unhindered outbreak. Across all outbreaks, 57% (90% CrI 47%– 66%) of all cases were estimated to have been averted (Fig. S3). Within each facility the impact of intervention varied, with the highest at location 2 with 79% (90% CrI 53% – 91%) cases averted and the lowest at location 11 with 9% (90% CrI −18% – 31%) cases averted. Under the model with multi-level intervention *ζ*, counterfactual modelling suggested a larger proportion of cases were averted: 73% (63%–78%) across all outbreaks.

**Figure 3:**
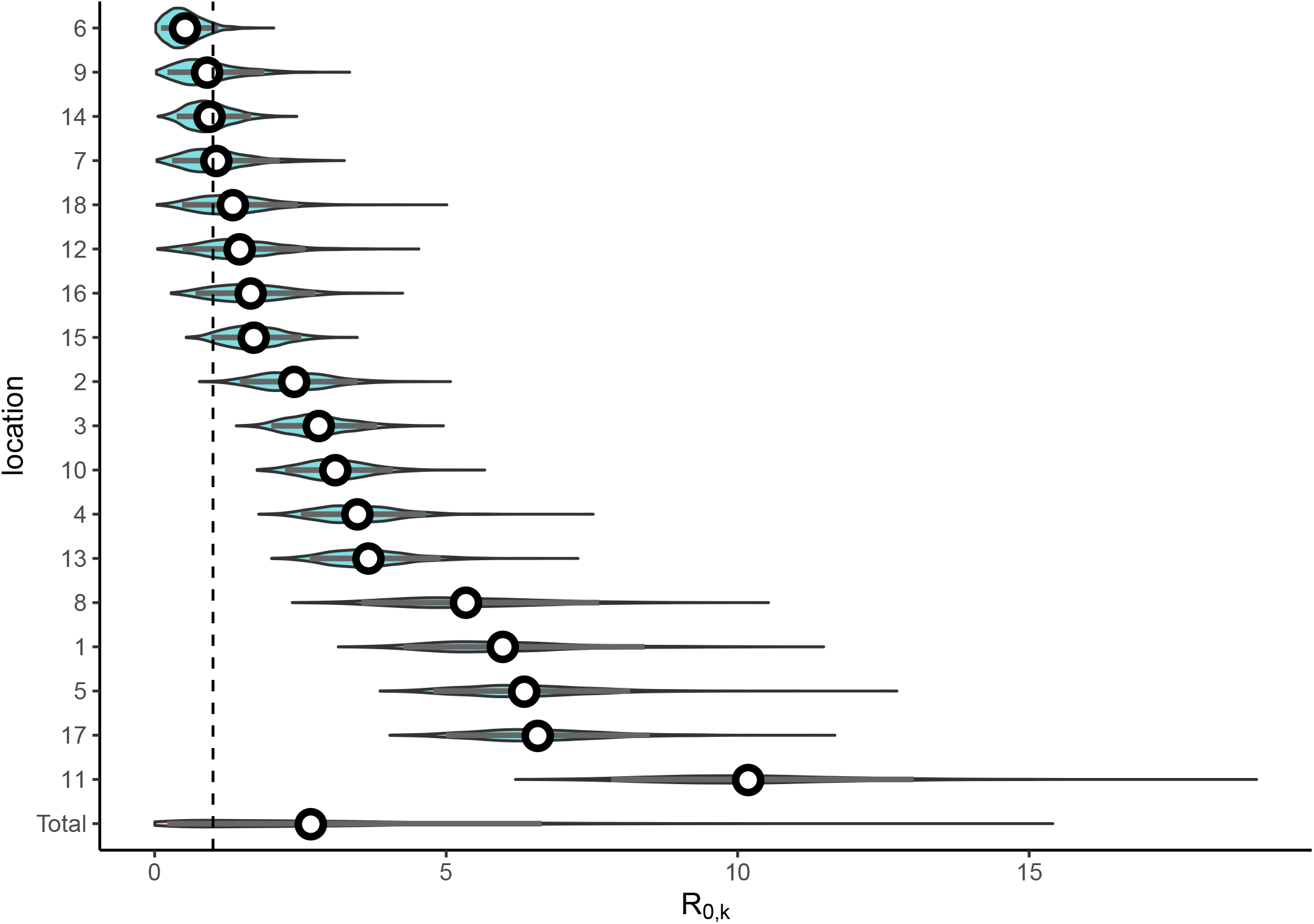
Hierarchical model fitting of *R*_0,*k*_. Each outbreak in a LTHC facility where the outbreak was greater than one individual was incorporated into the Bayesian hierarchical SEIR model that accounted for increased control measures once the first case had been identified. The posterior distribution of the *R*_0,*k*_ estimates are shown, with the posterior for *R*_0_ (Total) shown below. Median values are shown as white points, with the 90% credible interval as gray bars, the general distributions are shown as density violin plots.

**Figure 4:**
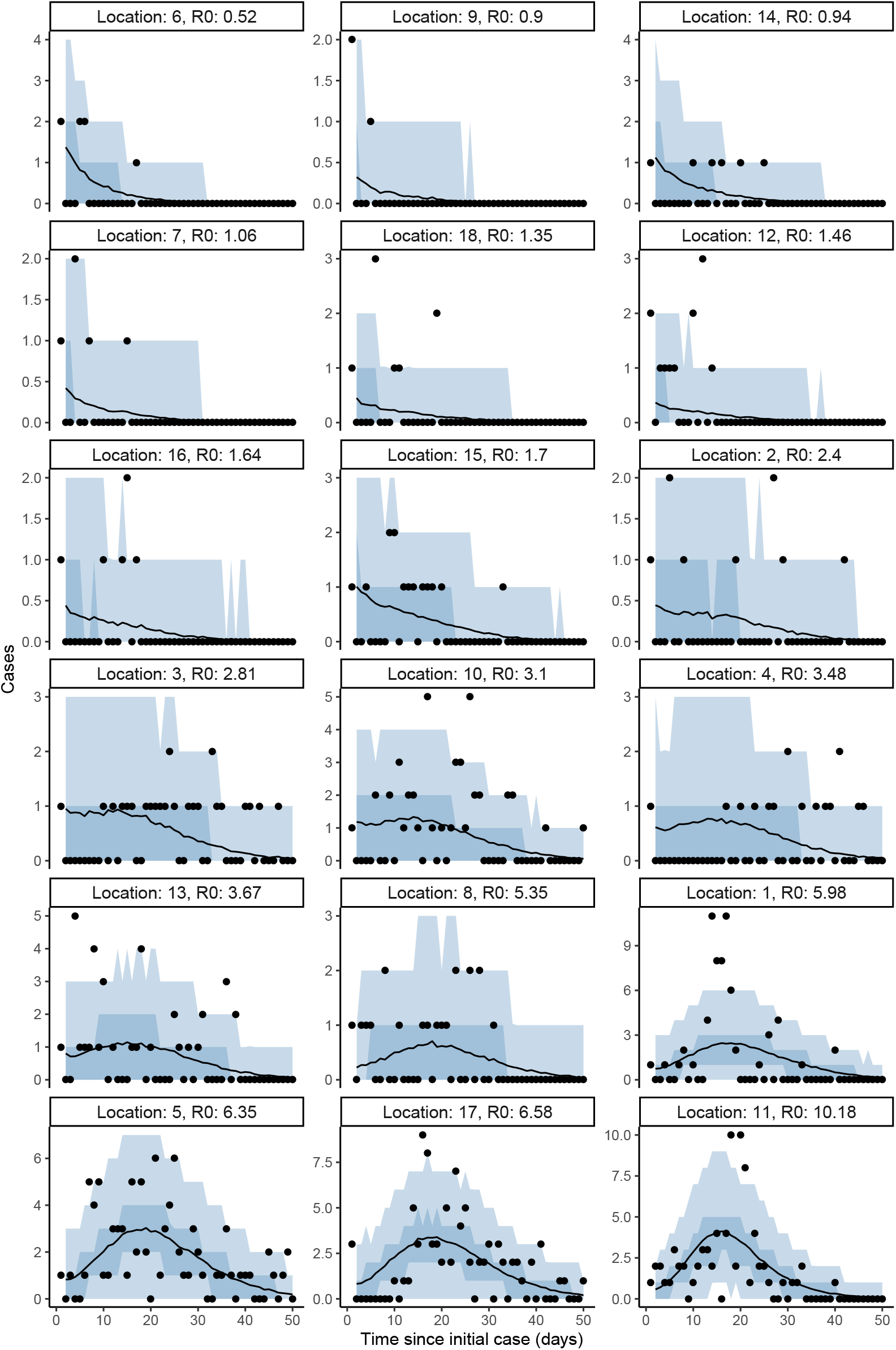
Bayesian hierarchical model fits to incident cases. For each LTHC outbreak of size greater than one, the posterior predictive distribution is shown. Median posterior values are shown as lines with the 50% and 90% credible intervals shown as darker and lighter shaded regions, respectively. Observed cases are overlaid as black points.

**Figure 5:**
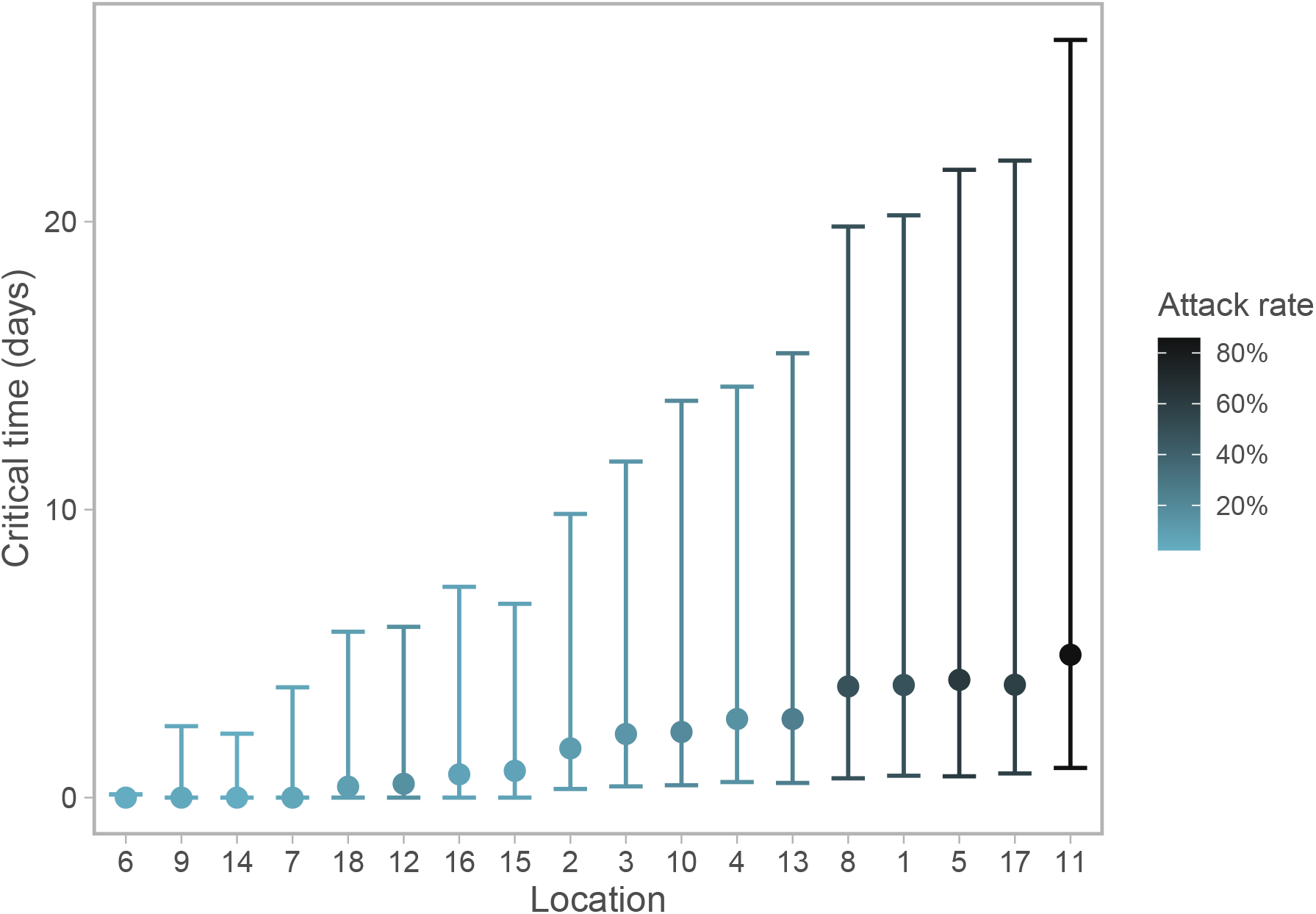
Posterior estimates of the critical time, from outbreak reported date to the reproduction number falling below 1. Calculated from posterior draws of *R*_0,*k*_ and *ζ*, with error bars showing 90% credible intervals.

The *R*_0,*k*_ estimates obtained from the EG and ML methods are quite different than those from the hierarchical model for many facilities (Fig. 6 & Table S2). Several of the smaller outbreaks (facilities 6, 7, 9, 15, 16) have a large degree of uncertainty around the EG and ML estimates, which is not seen in the estimates from the hierarchical model. For two outbreaks (facilities 4 and 7) the ML method was unable to obtain an estimate. There was limited concurrence between different methods as to the outbreaks with subcritical (< 1) *R*_0,*k*_ (EG: 2, 9, 12, 18; ML: 18; BHM: 6, 7, 9, 14). We see a general trend that for those outbreaks in which the hierarchical model estimated a smaller *R*_0,*k*_, the EG and ML methods, unguided by any prior or hierarchical information, have large levels of uncertainty (Fig. 6, ordered by increasing hierarchical model *R*_0,*k*_). For locations with larger *R*_0,*k*_ from the hierarchical model, we see consistently lower estimates from the EG and ML methods.

**Figure 6:**
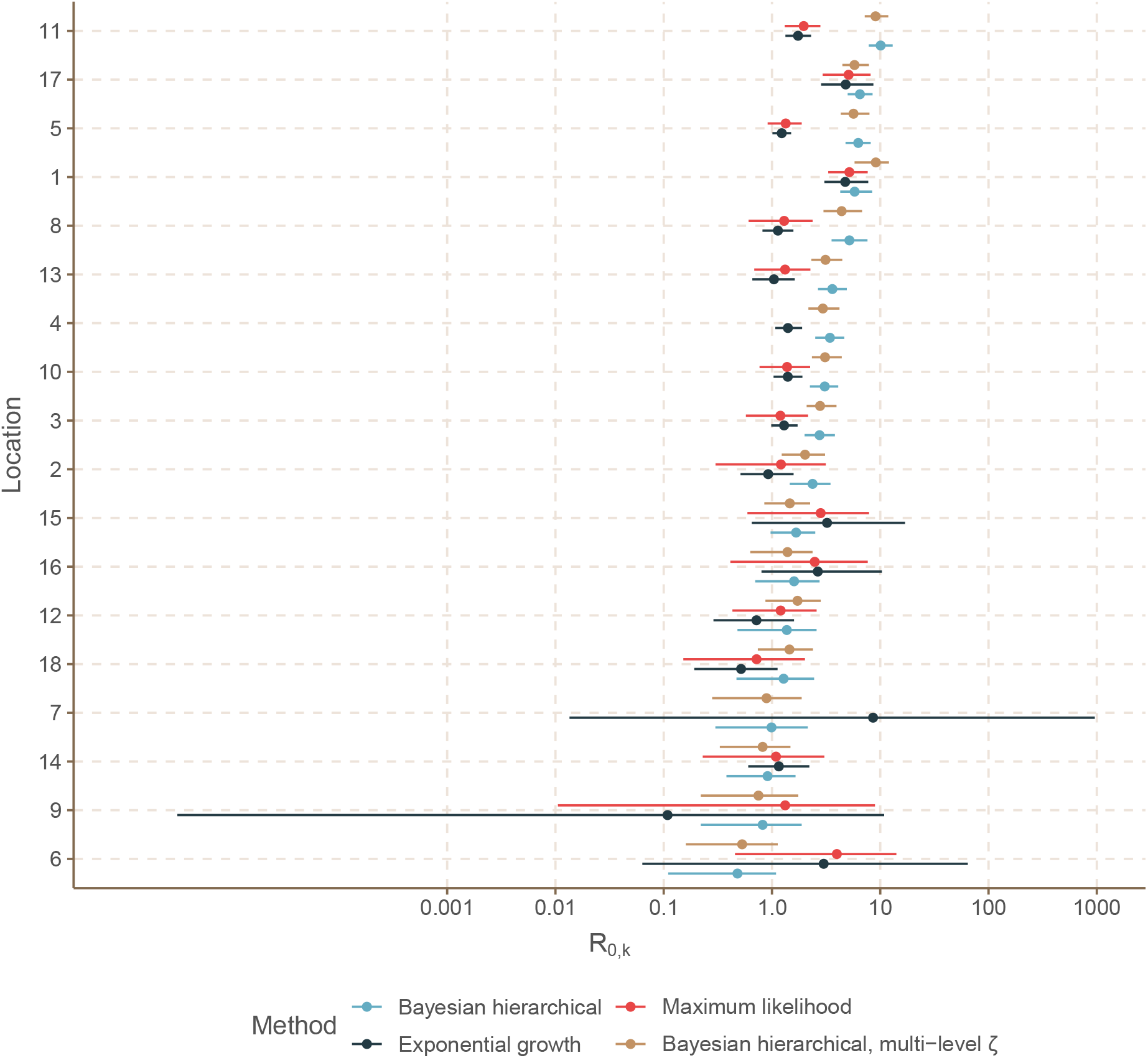
Comparison of *R*_0,*k*_ estimates (in log space) for COVID-19 outbreaks in LTHC facilities in BC, Canada. For each facility, *R*_0,*k*_ estimates with 95% confidence interval (exponential growth and maximum likelihood methods) or 90% credible interval (Bayesian hierarchical modelling, with and without multi-level *ζ* term) are shown.

To explore the impact of interpolating the missing symptom onset dates on the EG and ML *R*_0,*k*_ estimates, we used a Monte Carlo procedure for repeated random sampling of the missing dates. The resulting Monte Carlo means and confidence intervals can be found in Supplementary Table S1. We find that, despite the wide confidence intervals in Table S2 suggesting considerable uncertainty about the *R*_0,*k*_ estimates generally, the results are fairly robust to different interpolations of the missing symptom onset times (and many outbreaks have no missing symptom onset times). This would suggest that the majority of the uncertainty in the *R*_0,*k*_ estimates does not stem from the missing data, but from some other limitation(s) of the methods.

We include estimates of *R*_0,*k*_ directly from the attack rate in Table S2, though it is important to note that these are not directly comparable and do not truly represent ‘*R*_0_’ since they do not take into account the interventions put in place during the outbreak. Nonetheless, in the absence of temporal data this may be the only way to estimate *R*_0_, so we include these estimates for reference. There is, however, significant correlation (Pearson correlation coefficient PCC 0.98) between the *R*_0,*k*_ estimates from the Bayesian hierarchical modelling and the inferred attack rates (Figure S4), which is not found between the EG and ML *R*_0,*k*_ estimates and the attack rate (PCC 0.04 and 0.27).

We found only limited correlation between our estimates of *R*_0,*k*_ from the Bayesian hierarchical model and other factors regarding the LTHC facilities and their COVID-19 outbreaks, obtained from the Office of the Seniors Advocate, British Columbia [25] (Tables S3 and S4, Figure S5). No significant correlation was found between *R*_0,*k*_ and the initial case in a facility (by symptom onset date) being a staff member (PCC 0.09). For the continuous variables tested, all 95% confidence intervals on the correlation coefficient include zero, except for the number of lodged complaints during year 2018/19 for which we found a moderate negative correlation with *R*_0,*k*_ (PCC −0.53, p-value 0.04). Other small to moderate correlations were found between *R*_0,*k*_ and the average stay of residents in the LTHC facility (PCC 0.44), the number of direct care hours allocated per resident per day (PCC 0.39) and, to some extent, a negative correlation between *R*_0,*k*_ and the date on which the COVID-19 outbreak was reported in the facility (PCC −0.31, suggesting that later outbreaks were associated with a lower *R*_0,*k*_). However, p-values for these three tests are all above 0.05, and the sample size is relatively small; furthermore we were unable to obtain complete data on all facilities.

## 4. Discussion

In this paper we characterise variation in the estimated *R*_0_ within a series of COVID-19 outbreaks in LTHC facilities. As *R*_0_ is a product of infectiousness of the pathogen, contact rate, and type of contact, it is important to characterise it in order to understand the range of potential scenarios and total cases that could occur in future outbreaks, as well as subsequent capacity needs for healthcare provision, hospitals and intensive care units.

We found pronounced variability in *R*_0_ estimates by LTHC facility. Variation in *R*_0_ estimates could not be explained by facility size or whether the first infected individual was a healthcare worker. Small correlations were found between *R*_0_ and factors such as the average length of stay of facility residents (positive correlation) and the number of complaints lodged against the facility in 2018/19 (negative correlation), though the strength of these was not particularly significant. Further investigation of the covariate data would be required to more fully interpret these. Nonetheless, the majority of the variation in *R*_0_ estimates by facility was not found to be explainable by the factors considered. In small outbreaks stochastic effects can dominate, where random events can lead to a large number of secondary cases. It is important to note that these estimates do not explicitly model such events, but rather estimate the total secondary cases in aggregate. The variation in estimated *R*_0_ can then in part be explained by variation in these early events, between-person transmissibility, and other factors related to the individuals or facility. In addition, there is considerable evidence for the presence of super-spreader events and large heterogeneity in the number of secondary cases for SARS-CoV-2 [44, 45]. This heterogeneity can further increase the variance in *R*_0_ estimates both within and between outbreaks.

The Bayesian hierarchical model was able to regularize model fits even for outbreaks with a small number of cases, compared to the non-hierarchical methods for *R*_0_ estimation, which were independent between facilities and consequently suffered from high levels of uncertainty. This highlights a need for methods that consider totality of evidence. In model validation (Fig. S6), the hierarchical modelling procedure was able to recover the data-generating parameters. It is a flexible framework, which could be adapted for different types of outbreak outside of LTHC.

There are limitations to the study. The reported attack rate and modelled population size for LTHC facilities was based upon the maximum capacity for that facility, which may be larger than the total number of exposed individuals. Conversely, staff and family members may contribute to transmission in addition to facility residents. Although we allowed the maximum capacity size to vary to account for these factors, we lacked detailed data to inform this. Similarly, we did not consider a distinction between the behaviour of staff and residents. Although we estimated parameter *ζ*, the rate at which interventions brought *R*(*t*) below 1, the manner in which intervention curtailed transmission was not explicitly explored within this study, because the exact nature of interventions and their timings was not precisely known.

We considered all outbreaks as originating from a single index case in a closed population. Community prevalence remained low in BC during the period of this study so this seems a fair assumption, but it is possible that some facilities had multiple introductions. We also removed all outbreaks of size one since a single data point does not contain enough information to fit the compartmental model, but in leaving out those outbreaks we do lose some information about transmission. How much to account for this seems unclear, however, without further data on how much opportunity for transmission these single cases had. Although the fact that 30/53 LTHC outbreaks resulted in only 1 identified case certainly seems informative of the level of transmissibility within LTHC facilities, 25 of these were a single staff member infection (3 were resident cases, and 2 were not recorded). Without further data on these staff cases, we cannot know if an exposure event actually occurred within the LTHC facility.

The estimates of *R*_0_ in this work (from the initial period of each outbreak) do not represent the overall level of infectivity in the outbreaks, since interventions were swiftly put in place after the initial case was discovered. The attack rate was also low in many outbreaks, leading to potentially large effects of natural stochasticity on transmission. If trying to understand the potential for future outbreaks in LTHC, it is important to consider both *R*_0_ and *ζ* together. Importantly, we found that variation in *R*_0_ was not explainable by facility size or the initial case being a healthcare worker, but limited data meant we were also limited in what other covariates could be considered. Compared to some other jurisdictions, BC has also seen a relatively small number of large outbreaks. With sufficiently detailed data, this analysis could be strengthened by considering other geographical locations. Other studies have considered over-dispersion of secondary cases among outbreaks, finding high levels of over-dispersion among country-level outbreaks more generally [46]. It is not clear currently if this heterogeneity in secondary cases is due to contact patterns, age of cases, or other biological factors.

A counterfactual analysis of a scenario in which no interventions were put in place after the identification of an outbreak in each facility estimated that intervention led to 57% (47%–66%) of all cases being averted within LTHC facilities in BC (Fig. S3). This increased to 73% (63%–78%) in the model with hierarchical intervention *ζ*. This suggests that fast intervention after outbreak identification is key to controlling the spread of COVID-19. Estimates of the ‘critical time’ between implementation of interventions and the reproduction number dropping below one were all less than 5 days, however the credible intervals were often large, up to around 20 days. Particularly given the high fatality rates seen in LTHC facilities, implementing strong interventions rapidly will be particularly impactful in minimising morbidity and mortality. Facility outbreaks from later in the study period were weakly associated with lower critical times (PCC −0.31), suggesting some improvement in the implementation of control measures over time.

For future LTHC outbreaks, measures such as regular testing of staff could assist with identifying potential outbreaks as early as possible. Across the 18 LTHC outbreaks in this study, two thirds of those with known origin had a staff member as the initial case by symptom onset. At least 83% of those outbreaks with only 1 case had a staff member as the initial case. Prospective collection of data concerning factors that have been shown to impact spread of COVID-19 in other settings could also help reveal sources of variation in transmissibility between different LTHC settings: including ventilation systems, staff deployment practices, and patient layouts.

Despite evidence of improvement in the implementation of control measures in LTHC facilities over time, by the end of 2020 the number of reported long term care facility outbreaks in BC increased significantly to 224 [47]. This may largely be due to increased community prevalence [47], increasing the rate of importation to LTHC facilities, but also highlights significant remaining challenges in outbreak management. Increased community prevalence of COVID-19 would require a generalization of our current approach, which does not consider repeat importations to each LTHC facility, by estimation of a rate of case importation. This is a topic for future study, but could be highly informed by the current model in which we can more reasonably assume a single point of introduction. The recent increase in both community prevalence and the number of facility outbreaks in BC also highlights that LTHC facilities are strongly connected to their surrounding community, and this community plays a large role in protecting LTHC facilities from outbreaks, of COVID-19 and beyond. Although our findings suggest that the suite of interventions implemented in BC LTHC facilities has been effective in significantly reducing the size of COVID-19 outbreaks, the recent spike in outbreaks suggests that further improving these controls is necessary to reduce disease burden and associated mortality in LTHC facilities under increased community transmission.

## 5. Data, code and materials

The methodology described in this work for Bayesian hierarchical SEIR fitting to a collection of outbreak data from different facilities is available as an R package at github.com/sempwn/cr0eso [48]. This package also contains code and de-identified data to reproduce the analysis in this manuscript. Covariate LTHC facility data was obtained from seniorsadvocatebc.ca/quickfacts [25]. Additional cumulative data on LTHC outbreaks in BC is available from bccdc.ca/health-info/diseases-conditions/covid-19/data [49].

## Data Availability

The methodology described in this work for Bayesian hierarchical SEIR fitting to a collection of outbreak data from different facilities is available as an R package at github.com/sempwn/cr0eso. This package also contains code and de-identified data to reproduce the analysis in this manuscript. Covariate LTHC facility data was obtained from seniorsadvocatebc.ca/quickfacts. Additional cumulative data on LTHC outbreaks in BC is available from bccdc.ca/healthinfo/
diseases-conditions/covid-19/data.

https://www.seniorsadvocatebc.ca/quickfacts

http://www.bccdc.ca/health-info/diseases-conditions/covid-19/data

## 8. Authors’ Contributions

Conceptualization and discussion: all authors. Data acquisition and curation: JES, MCO, NZJ, MAI. Bayesian model development: MAI. Analysis and visualization: JES, MAI. Software: JES, SCA, MAI. Writing – original draft: JES, MAI. Writing – review, editing: all authors. All authors gave final approval for publication and agree to be held accountable for the work performed therein.

## 9. Acknowledgements

The authors thank Dr. Rohit Vijh for their advice and perspectives, and Fisheries and Oceans Canada for their support. Thank you to the British Columbia Centre for Disease Control, Fraser Health Authority and Vancouver Coastal Health Authority for sharing the data used in this analysis.

## References

[1] Shen Y, Li C, Dong H, Wang Z, Martinez L, Sun Z, et al. Community outbreak investigation of SARS-CoV-2 transmission among bus riders in eastern China. JAMA Intern Med 2020;180(12):1665–71. doi:10.1001/jamainternmed.2020.5225.

[2] Hamner L, Dubbel P, Capron I, Ross A, Jordan A, Lee J, et al. High SARS-CoV-2 Attack Rate Following Exposure at a Choir Practice—Skagit County, Washington, March 2020. MMWR Morb Mortal Wkly Rep 2020;69:606–10. doi:10.15585/mmwr.mm6919e6.

[3] Zhang S, Diao M, Yu W, Pei L, Lin Z, Chen D. Estimation of the reproductive number of novel coronavirus (COVID-19) and the probable outbreak size on the Diamond Princess cruise ship: A data-driven analysis. Int J Infect Dis 2020;93:201–4. doi:10.1016/j.ijid.2020.02.033.

[4] Dyal JW, Grant MP, Broadwater K, Bjork A, Waltenburg MA, Gibbins JD, et al. COVID-19 Among Workers in Meat and Poultry Processing Facilities—19 States, April 2020. MMWR Morbidity and Mortality Weekly Report 2020;69:557–61. doi:10.15585/mmwr.mm6918e3.

[5] Danis K, Epaulard O, Bénet T, Gaymard A, Campoy S, Bothelo-Nevers E, et al. Cluster of coronavirus disease 2019 (COVID-19) in the French Alps, 2020. Clinical Infectious Diseases 2020;71(15):825–32. doi:10.1093/cid/ciaa424.

[6] Wallace M, Hagan L, Curran KG, Williams SP, Handanagic S, Bjork SL, et al. COVID-19 in correctional and detention facilities—United States, February–April 2020. MMWR Morbidity and mortality weekly report 2020;69:587–90. doi:10.15585/mmwr.mm6919e1.

[7] Addetia A, Crawford KHD, Dingens A, Zhu H, Roychoudhury P, Huang M, et al. Neutralizing antibodies correlate with protection from SARS-CoV-2 in humans during a fishery vessel outbreak with a high attack rate. J Clin Microbiol 2020;58(11):e02107–20. doi:10.1128/JCM.02107-20.

[8] Kinner SA, Young JT, Snow K, Southalan L, Lopez-Acuña D, Ferreira-Borges C, et al. Prisons and custodial settings are part of a comprehensive response to COVID-19. Lancet Public Health 2020;5(4):e188–9. doi:10.1016/S2468-2667(20)30058-X.

[9] Irvine M, Coombs D, Skarha J, del Pozo B, Rich J, Taxman F, et al. Modeling COVID-19 and its impacts on US Immigration and Customs Enforcement (ICE) detention facilities, 2020. J Urban Health 2020;97:439–447. doi:10.1007/s11524-020-00441-x.

[10] Van Houtven CH, Boucher NA, Dawson WD. Impact of the COVID-19 outbreak on long-term care in the United States. International Long-Term Care Policy Network, CPEC-LSE 2020;URL: https://ltccovid.org.

[11] Comas-Herrera A, Zalakaín J, Litwin C, Hsu AT, Lane N, Fernández JL. Mortality associated with COVID-19 outbreaks in care homes: early international evidence. International Long-Term Care Policy Network, CPEC-LSE 2020;URL: https://ltccovid.org.

[12] McMichael TM, Currie DW, Clark S, Pogosjans S, Kay M, Schwartz NG, et al. Epidemiology of COVID-19 in a long-term care facility in King County, Washington. N Engl J Med 2020;382(21):2005–11. doi:10.1056/NEJMoa2005412.

[13] Hsu AT, Lane N. Impact of COVID-19 on residents of Canada’s long-term care homes– ongoing challenges and policy response. International Long-Term Care Policy Network, CPEC-LSE 2020;URL: https://ltccovid.org.

[14] Anderson SC, Mulberry N, Edwards AM, Stockdale JE, Iyaniwura SA, Falcao RC, et al. How much leeway is there to relax covid-19 control measures? medRxiv preprint 2020;doi:10.1101/2020.06.12.20129833.

[15] Liu Y, Gayle AA, Wilder-Smith A, Rocklöv J. The reproductive number of COVID-19 is higher compared to SARS coronavirus. J Travel Med 2020;27(2). doi:10.1093/jtm/taaa021.

[16] Hilton J, Keeling MJ. Estimation of country-level basic reproductive ratios for novel Coronavirus (SARS-CoV-2/COVID-19) using synthetic contact matrices. PLoS Comput Biol 2020;16(7):e1008031. doi:10.1371/journal.pcbi.1008031.

[17] Tuite AR, Fisman DN, Greer AL. Mathematical modelling of COVID-19 transmission and mitigation strategies in the population of Ontario, Canada. CMAJ 2020;192(19):E497–505. doi:10.1503/cmaj.200476.

[18] Donnat C, Holmes S. Modeling the heterogeneity in COVID-19’s reproductive mumber and its impact on predictive scenarios. arXiv preprint 2020;URL: https://arxiv.org/abs/2004.05272.

[19] Logar S. Care home facilities as new COVID-19 hotspots: Lombardy Region (Italy) case study. Arch Gerontol Geriatr 2020;89:104087. doi:10.1016/j.archger.2020.104087.

[20] Stall NM, Jones A, Brown KA, Rochon PA, Costa AP. For-profit long-term care homes and the risk of COVID-19 outbreaks and resident deaths. CMAJ 2020;192(33):E946–55. doi:10.1503/cmaj.201197.

[21] Burton JK, Bayne G, Evans C, Garbe F, Gorman D, Honhold N, et al. Evolution and effects of COVID-19 outbreaks in care homes: a population analysis in 189 care homes in one geographical region of the UK. Lancet Healthy Longevity 2020;1(1). doi:10.1016/S2666-7568(20)30012-X.

[22] Béland D, Marier P. COVID-19 and long-term care policy for older people in Canada. J Aging Soc Policy 2020;32(4–5):358–64. doi:10.1080/08959420.2020.1764319.

[23] BC Centre for Disease Control and BC Ministry of Health. Infection Prevention and Control Requirements for COVID-19 in Long Term Care and Seniors’ Assisted Living. http://www.bccdc.ca/Health-Info-Site/Documents/COVID19_LongTermCareAssistedLiving.pdf; 2020. Accessed: July 6th, 2020.

[24] Vijh R, Prairie J, Otterstatter M, Hu Y, Hayden A, Yau B, et al. Evaluation of a multisectoral intervention to mitigate the risk of sars-cov-2 transmission in long-term care facilities. Infection Control & Hospital Epidemiology 2021;:1–37doi:10.1017/ice.2020.1407.

[25] Office of the Seniors Advocate, British Columbia. Long-Term Care Facilities Quick Facts Directory. https://www.seniorsadvocatebc.ca/quickfacts/location; 2020. Accessed: November 30th, 2020.

[26] Hsu C, Yen A, Chen L, Chen H. Analysis of household data on influenza epidemic with Bayesian hierarchical model. Math Biosci 2015;261:13–26. doi:10.1016/j.mbs.2014.11.006.

[27] Comin A, Klinkenberg D, Stegeman A, Busani L, Marangon S, et al. Estimate of basic reproduction number (R_0_) of low pathogenicity avian influenza outbreaks using a bayesian approach. Society for Veterinary Epidemiology and Preventive Medicine Proceedings, Nantes, France, 24-26 March, 2010 2010;:145–53.

[28] Osthus D, Gattiker J, Priedhorsky R, Del Valle SY, et al. Dynamic Bayesian influenza forecasting in the United States with hierarchical discrepancy (with discussion). Bayesian Analysis 2019;14(1):261–312. doi:10.1214/18-BA1117.

[29] Gelman A, Hill J. Data analysis using regression and multilevel/hierarchical models. Cambridge University Press; 2006.

[30] McAloon C, Collins Á, Hunt K, Barber A, Byrne AW, Butler F, et al. Incubation period of COVID-19: a rapid systematic review and meta-analysis of observational research. BMJ open 2020;10(8):e039652. doi:10.1136/bmjopen-2020-039652.

[31] Alimohamadi Y, Taghdir M, Sepandi M. The estimate of the basic reproduction number for novel coronavirus disease (COVID-19): a systematic review and meta-analysis. J Prev Med Public Health 2020;53(3):151–7. doi:10.3961/jpmph.20.076.

[32] Hoffman MD, Gelman A. The No-U-Turn sampler: adaptively setting path lengths in Hamiltonian Monte Carlo. J Mach Learn Res 2014;15(1):1593–623.

[33] Gelman A, Carlin JB, Stern HS, Dunson DB, Vehtari A, Rubin DB. Bayesian data analysis. CRC press; 2013.

[34] Stan Development Team. RStan: the R interface to Stan. 2020. URL: http://mc-stan.org/; R package version 2.21.2.

[35] Lessler J, Cummings DA, Fishman S, Vora A, Burke DS. Transmissibility of swine flu at Fort Dix, 1976. J R Soc Interface 2007;4(15):755–62. doi:10.1098/rsif.2007.0228.

[36] Diekmann O, Heesterbeek JAP. Mathematical epidemiology of infectious diseases: model building, analysis and interpretation. Wiley; 1989.

[37] Dietz K. The estimation of the basic reproduction number for infectious diseases. Stat Methods Med Res 1993;2(1):23–41. doi:10.1177/096228029300200103.

[38] Wallinga J, Lipsitch M. How generation intervals shape the relationship between growth rates and reproductive numbers. Proc Royal Soc B 2007;274:599–604. doi:10.1098/rspb.2006.3754.

[39] White LF, Wallinga J, Finelli L, Reed C, Riley S, Lipsitch M, et al. Estimation of the reproductive number and the serial interval in early phase of the 2009 influenza A/H1N1 pandemic in the USA. Influenza Other Respir Viruses 2009;3(6):267–76. doi:10.1111/j.1750-2659.2009.00106.x.

[40] Ganyani T, Kremer C, Chen D, Torneri A, Faes C, Wallinga J, et al. Estimating the generation interval for coronavirus disease (COVID-19) based on symptom onset data, March 2020. Eurosurveillance 2020;25(17):2000257. doi:10.2807/1560-7917.ES.2020.25.17.2000257.

[41] Obadia T, Haneef R, Boëlle PY. The R0 package: a toolbox to estimate reproduction numbers for epidemic outbreaks. BMC Med Inform Decis Mak 2012;12(147). doi:10.1186/1472-6947-12-147.

[42] Wickham H, Averick M, Bryan J, Chang W, McGowan LD, François R, et al. Welcome to the tidyverse. J Open Source Softw 2019;4(43):1686. doi:10.21105/joss.01686.

[43] R Core Team. R: A Language and Environment for Statistical Computing. R Foundation for Statistical Computing; Vienna, Austria; 2020. URL: https://www.R-project.org/.

[44] Zhang Y, Li Y, Wang L, Li M, Zhou X. Evaluating transmission heterogeneity and super-spreading event of COVID-19 in a metropolis of China. Int J Environ Res Public Health 2020;17(10):3705. doi:10.3390/ijerph17103705.

[45] Wang L, Didelot X, Yang J, Wong G, Shi Y, Liu W, et al. Inference of person-to-person transmission of covid-19 reveals hidden super-spreading events during the early outbreak phase. Nat Commun 2020;11:5006. doi:10.1038/s41467-020-18836-4.

[46] Endo A, Abbott S, Kucharski AJ, Funk S, et al. Estimating the overdispersion in covid-19 transmission using outbreak sizes outside china. Wellcome Open Res 2020;5(67). doi:10.12688/wellcomeopenres.15842.3.

[47] BC Centre for Disease Control. British Columbia (BC) COVID-19 Situation Report Week 50: December 6 – December 12, 2020. http://www.bccdc.ca/Health-Info-Site/Documents/COVID_sitrep/Week_50_BC_COVID-19_Situation_Report.pdf; 2020. Accessed: January 6th, 2021.

[48] Covid R0 estimation of outbreaks (cr0eso). https://github.com/sempwn/cr0eso; 2020.

[49] BC Centre for Disease Control. BC COVID-19 Data. http://www.bccdc.ca/health-info/diseases-conditions/covid-19/data ; 2020. Accessed: January 26th, 2021.

