## Supplemental Materials for "Quantifying transmissibility of COVID-19 and impact of intervention within long-term health care facilities"

| Location | $n$ | No. missing symptom onsets | $R_{0,k}$, EG Monte Carlo mean (95% CI) | $R_{0,k}$, ML Monte Carlo mean (95% CI) |
|---|---|---|---|---|
| 1 | 67 | 8 | 5.20 (3.02–7.38) | 4.92 (3.69–6.15) |
| 2 | 9 | 0 | 0.92 (0.92–0.92) | 1.21 (1.21–1.21) |
| 3 | 25 | 0 | 1.29 (1.29–1.29) | 1.20 (1.20–1.20) |
| 4 | 18 | 0 | 1.40 (1.40–1.40) | – |
| 5 | 88 | 6 | 1.73 (0.0–6.97) | 1.59 (0.0–3.82) |
| 6 | 7 | 3 | 10.63 (0.0–30.11) | 5.46 (0.0–11.95) |
| 7 | 5 | 0 | 8.58 (8.58–8.58) | – |
| 8 | 19 | 0 | 1.13 (1.13–1.13) | 1.30 (1.30–1.30) |
| 9 | 3 | 0 | 0.11 (0.11–0.11) | 1.32 (1.32–1.32) |
| 10 | 44 | 13 | 2.33 (0.0–4.73) | 2.02 (0.57–3.47) |
| 11 | 79 | 0 | 1.74 (1.74–1.74) | 1.96 (1.96–1.96) |
| 12 | 12 | 3 | 1.00 (0.0–2.84) | 1.87 (0.0–5.97) |
| 13 | 37 | 2 | 18.79 (0.0–97.19) | 2.42 (0.0–7.40) |
| 14 | 6 | 0 | 1.16 (1.16–1.16) | 1.09 (1.09–1.09) |
| 15 | 14 | 0 | 3.23 (3.23–3.23) | 2.82 (2.82–2.82) |
| 16 | 6 | 0 | 2.65 (2.65–2.65) | 2.48 (2.48–2.48) |
| 17 | 89 | 7 | 4.98 (3.29–6.68) | 5.34 (3.42–7.27) |
| 18 | 8 | 2 | 6.65 (0.0–19.73) | 3.19 (0.0–6.87) |

Table S1: **Results of sensitivity analysis of interpolated symptom onset dates on the EG and ML $R_{0,k}$ estimates**. 100 sets of missing onset times were sampled, and $R_{0,k}$ estimates calculated for each. Shown are the mean and 95% confidence intervals of the 100 $R_{0,k}$ estimates.

| Facility | Reported date | $n$ | $N$ | Attack rate (%) | $R_{0,k}$ ($A_r$) | $R_{0,k}$ (EG) | $R_{0,k}$ (ML) | $R_{0,k}$ (BHM) | $R_{0,k}$ (multi-level intervention) | $\zeta$ (multi-level intervention) | Critical time (BHM) |
|---|---|---|---|---|---|---|---|---|---|---|---|
| 1 | 2020-04-28 | 67 | 139 | 48.2 (41.9–56.7) | 1.36 (1.29–1.46) | 4.74 (3.05–7.75) | 5.18 (3.31–7.66) | 5.8 (4.27–8.41) | 9.1 (5.79–12.03) | 0.43 (0.07–2.19) | 3.91 (0.76–20.22) |
| 2 | 2020-04-25 | 9 | 91 | 9.9 (8.6–11.6) | 1.05 (1.04–1.06) | 0.92 (0.51–1.58) | 1.21 (0.30–3.14) | 2.37 (1.46–3.47) | 2.02 (1.23–3.09) | 0.4 (0.07–2.07) | 1.71 (0.3–9.85) |
| 3 | 2020-03-16 | 25 | 189 | 13.2 (11.5–15.6) | 1.07 (1.06–1.08) | 1.29 (0.98–1.72) | 1.20 (0.57–2.15) | 2.75 (2–3.81) | 2.78 (2.09–3.94) | 0.38 (0.07–2.17) | 2.21 (0.39–11.67) |
| 4 | 2020-03-22 | 18 | 120 | 15.0 (13.0–17.6) | 1.08 (1.07–1.10) | 1.40 (1.07–1.90) | – | 3.43 (2.51–4.65) | 2.95 (2.17–4.2) | 0.38 (0.07–2.17) | 2.73 (0.54–14.27) |
| 5 | 2020-06-09 | 88 | 142 | 62.0 (53.9–72.9) | 1.56 (1.42–1.75) | 1.23 (1.01–1.50) | 1.34 (0.91–1.88) | 6.26 (4.78–8.16) | 5.66 (4.32–7.95) | 0.41 (0.06–1.93) | 4.09 (0.74–21.8) |
| 6 | 2020-08-06 | 7 | 300 | 2.3 (2.0–2.7) | 1.01 (1.01–1.01) | 3.00 (0.06–64.35) | 3.97 (0.46–14.09) | 0.48 (0.08–2.36) | 0.53 (0.16–1.13) | 0.39 (0.06–2.04) | 0.0 (0.0–0.11) |
| 7 | 2020-04-28 | 5 | 87 | 5.7 (5–6.8) | 1.03 (1.03–1.03) | 8.58 (0.01–958.23) | – | 0.99 (0.3–2.14) | 0.89 (0.28–1.88) | 0.4 (0.06–2.06) | 0.0 (0.0–3.83) |
| 8 | 2020-03-29 | 19 | 40 | 47.5 (41.3–55.9) | 1.36 (1.28–1.45) | 1.13 (0.82–1.58) | 1.30 (0.61–2.37) | 5.19 (3.55–7.62) | 4.4 (2.99–6.79) | 0.41 (0.05–2.17) | 3.86 (0.67–19.83) |
| 9 | 2020-03-25 | 3 | 66 | 4.5 (4.0–5.3) | 1.02 (1.02–1.03) | 0.11 (0.0–10.86) | 1.32 (0.01–8.94) | 0.82 (0.22–1.88) | 0.75 (0.22–1.75) | 0.36 (0.07–2.26) | 0.0 (0.0–2.48) |
| 10 | 2020-08-08 | 44 | 236 | 18.6 (16.2–21.9) | 1.11 (1.09–1.13) | 1.40 (1.03–1.91) | 1.38 (0.77–2.25) | 3.07 (2.24–4.09) | 3.09 (2.34–4.41) | 0.39 (0.07–2.26) | 2.28 (0.43–13.78) |
| 11 | 2020-03-05 | 79 | 92 | 85.9 (74.7–101) | 2.28 (1.79–4.44) | 1.74 (1.33–2.30) | 1.96 (1.31–2.80) | 10.08 (7.83–13.02) | 9.04 (7.19–11.86) | 0.44 (0.07–1.9) | 4.96 (1.03–26.31) |
| 12 | 2020-09-02 | 12 | 75 | 16.0 (13.9–18.8) | 1.09 (1.07–1.11)) | 0.72 (0.29–1.60) | 1.20 (0.43-2.58) | 1.37 (0.48–2.58) | 1.72 (0.87–2.82) | 0.4 (0.07–2) | 0.48 (0.0–5.93) |
| 13 | 2020-03-28 | 37 | 151 | 24.5 (21.3–28.8) | 1.15 (1.12–1.17) | 1.04 (0.66–1.62) | 1.32 (0.69-2.26) | 3.61 (2.66–4.91) | 3.12 (2.31–4.46) | 0.36 (0.06–1.99) | 2.73 (0.51–15.43) |
| 14 | 2020-04-22 | 6 | 236 | 2.5 (2.2–3.0) | 1.01 (1.01–1.01) | 1.16 (0.60–2.21) | 1.09 (0.23–3.05) | 0.91 (0.38–1.65) | 0.82 (0.33–1.48) | 0.42 (0.07–2.28) | 0.0 (0.0–2.22) |
| 15 | 2020-03-26 | 14 | 210 | 6.7 (5.8–7.8) | 1.03 (1.03–1.04) | 3.23 (0.65–16.93) | 2.82 (0.59–7.88) | 1.67 (0.97–2.51) | 1.46 (0.85–2.25) | 0.42 (0.07–2.15) | 0.93 (0.0–6.73) |
| 16 | 2020-03-26 | 6 | 92 | 6.5 (5.7–7.7) | 1.03 (1.03–1.04) | 2.65 (0.80–10.35) | 2.48 (0.41–7.66) | 1.6 (0.7–2.75) | 1.39 (0.63–2.37) | 0.41 (0.07–2.12) | 0.81 (0.0–7.32) |
| 17 | 2020-03-17 | 89 | 154 | 57.8 (50.3–68.0) | 1.49 (1.38–1.64) | 4.79 (2.84–8.64) | 5.10 (2.94–8.14) | 6.49 (5–8.49) | 5.78 (4.47–7.86) | 0.38 (0.07–2.19) | 3.92 (0.84–22.12) |
| 18 | 2020-04-21 | 8 | 89 | 9.0 (7.8–10.6) | 1.05 (1.04–1.06) | 0.52 (0.19–1.13) | 0.72 (0.15–2.02) | 1.28 (0.47–2.45) | 1.45 (0.74–2.39) | 0.4 (0.06–2.14) | 0.38 (0.0–5.76) |
| Total | – | – | – | – | – | – | – | 2.19 (0.19 - 6.69) | 2.86 (0.35 - 7.97) | 0.4 (0.07–2.15) | – |

Table S2: **Results of $R_0$ estimation within LTHC outbreaks in BC, Canada**. Attack rate ($A_r$) and corresponding $R_{0,k}$ were estimated from the final outbreak size and the maximum capacity of the facility: capacity was varied between 85%–115% to produce the reported ranges. Exponential growth (EG) and maximum likelihood (ML) $R_{0,k}$ estimates shown with 95% confidence intervals. For two of the outbreaks, the ML method was not able to obtain an estimate. $R_{0,k}$ estimates obtained from the Bayesian hierarchical model (BHM) shown with 90% credible intervals, and again with inclusion of multi-level intervention $\zeta$. The critical time (with 90% credible intervals) is calculated as the number of days taken after interventions were implemented for $R_k(t)$ to fall below 1 in the hierarchical model with fixed intervention. 'Total' estimates are taken from the predictive distribution.

| Factor | Correlation (95% CI) |
|---|---|
| Average resident age (years) | -0.02 (-0.53, 0.5) |
| Average resident stay (days) | 0.44 (-0.1, 0.78) |
| COVID-19 outbreak reported date | -0.31 (-0.68, 0.19) |
| Direct care hours /resident/day | 0.39 (-0.15, 0.75) |
| Facility capacity | -0.21 (-0.62, 0.29) |
| Number of disease outbreaks 2018/19 | -0.18 (-0.64, 0.36) |
| Number of lodged complaints 2018/19 | -0.53 (-0.82, -0.03) |
| Residents dependent for daily activities (%) | 0.16 (-0.39, 0.62) |
| Year facility opened | -0.16 (-0.64, 0.40) |

Table S3: **Correlation between $R_{0,k}$ estimates from Bayesian hierarchical modelling and various LTHC facility factors, using Pearson's product moment correlation coefficient.** All factor data other than COVID-19 outbreak reported date and facility capacity is from the 2018/19 year [S1].

| Factor | Correlation |
|---|---|
| Accreditation status | 0.25 |
| Identity of initial COVID-19 case | 0.09 |

Table S4: **Point-biserial (Pearson) correlation between $R_{0,k}$ estimates from Bayesian hierarchical modelling and dichotomous categorical LTHC facility factors** Accreditation status data is from the 2018/19 year [S1]. A positive correlation would correspond to a positive relationship between $R_0$ and a facility being accredited or the initial case being a staff member, respectively.

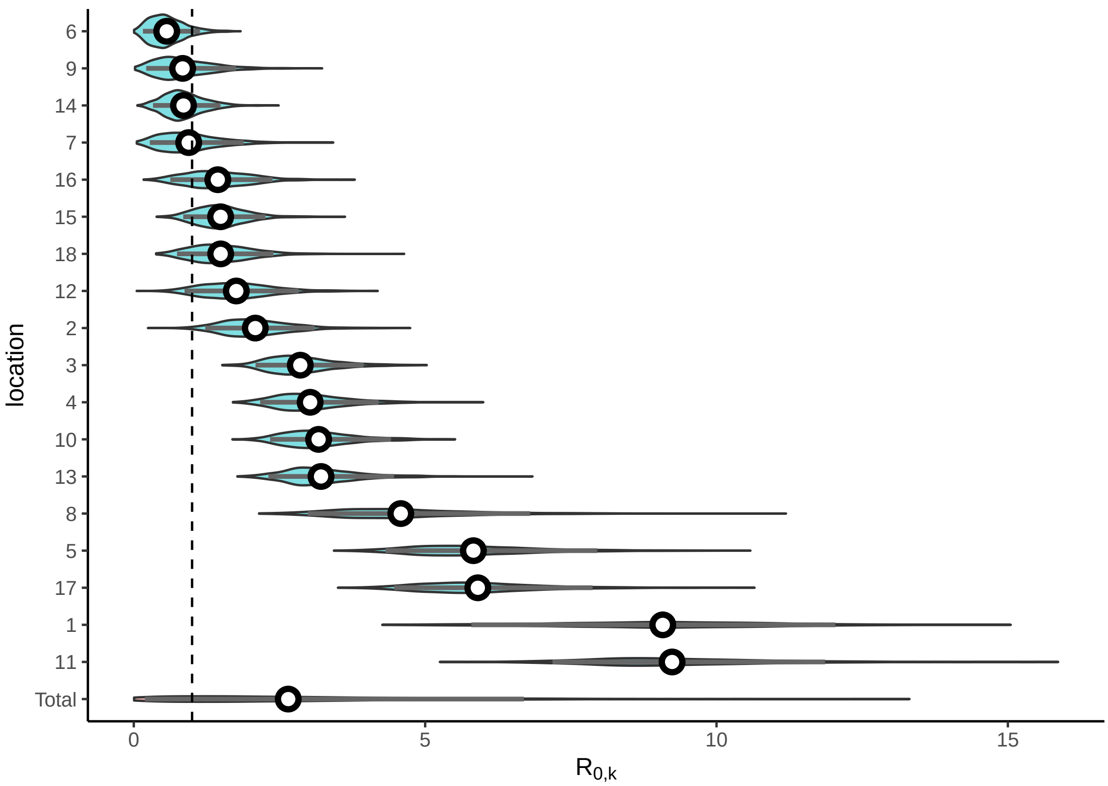


(a) Multi-level $R_{0,k}$ marginal posterior distributions

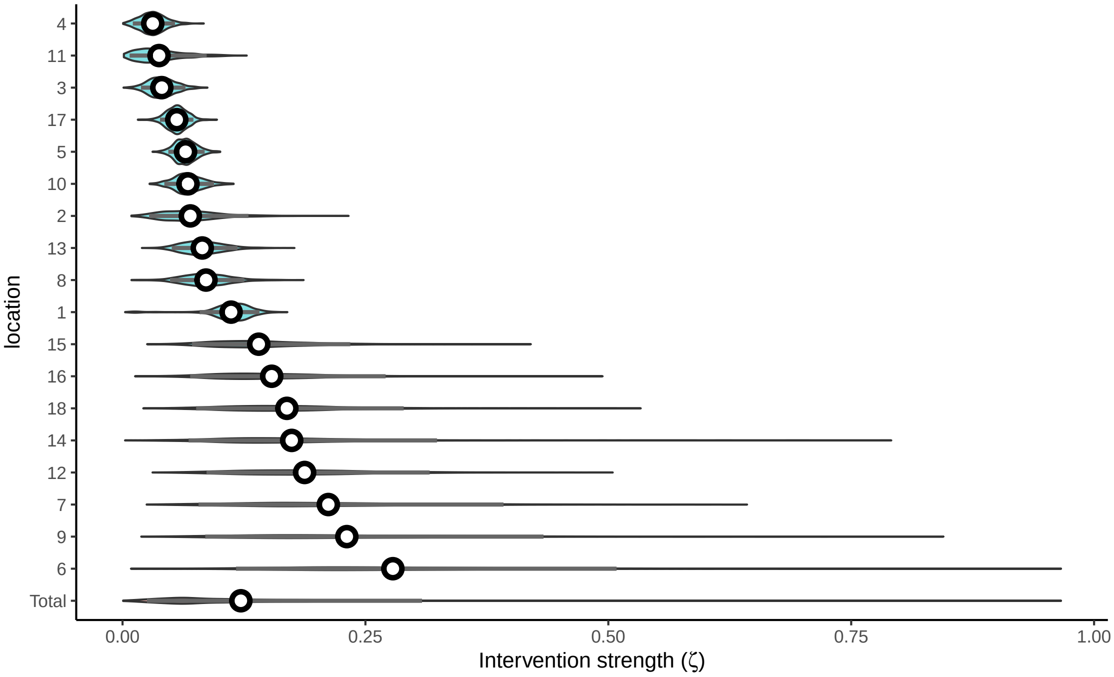


(b) Multi-level $\zeta$ marginal posterior distributions

Figure S1: **Hierarchical model fitting of $R_0$ with multi-level $R_{0,k}$ and $\zeta$ terms.** Posterior predictive draws of $R_{0,k}$ and $\zeta$ are shown, with median values (white points), 90% credible intervals (gray bars) and general distribution (violin plots).

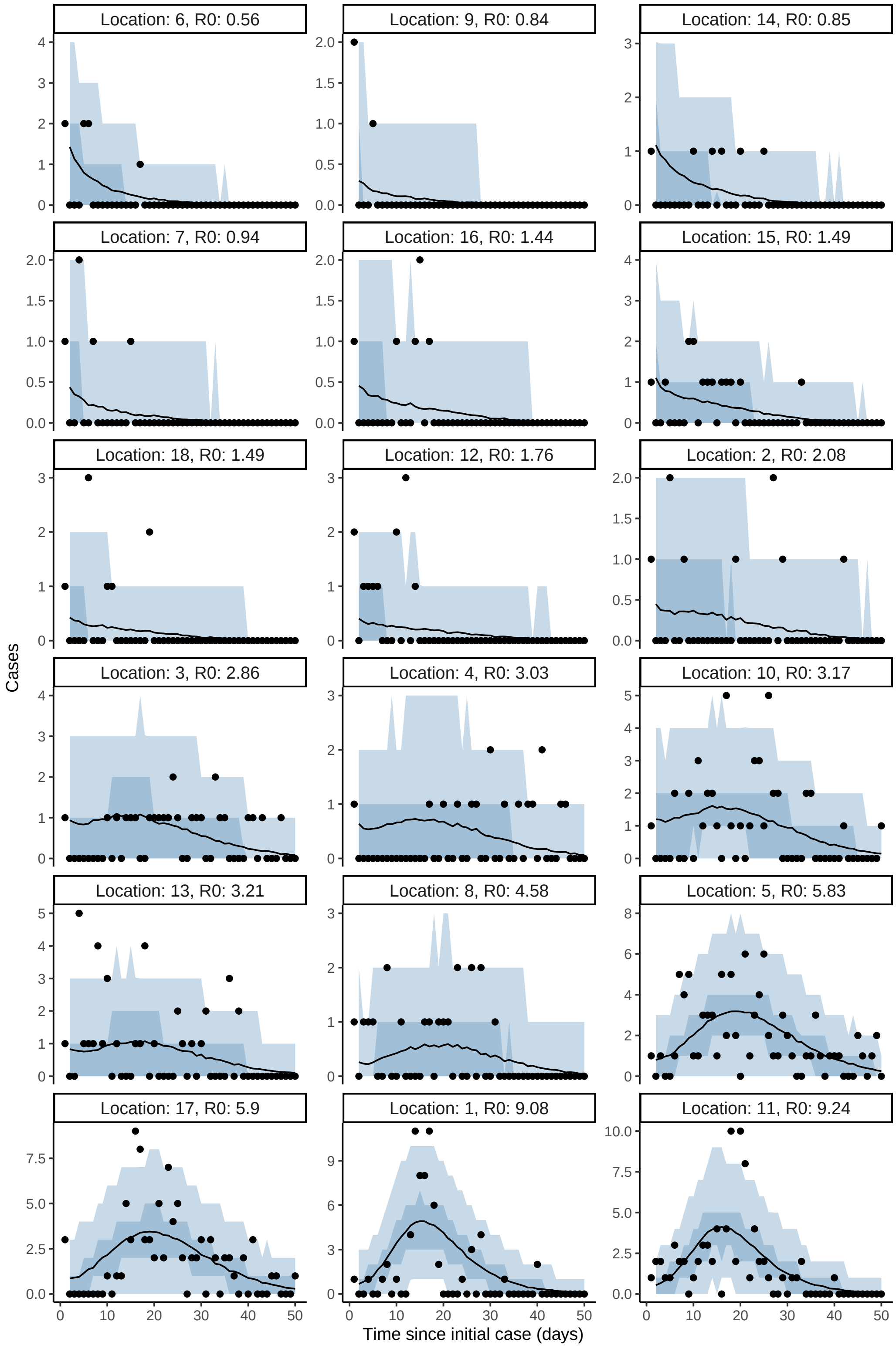


Figure S2: **Model fit to incidence within each facility for hierarchical $R_{0,k}$ and $\zeta$ model.** Median posterior values are shown as lines with the 50% and 90% credible intervals as darker and lighter shaded regions, respectively. Observed cases are overlaid as black points.

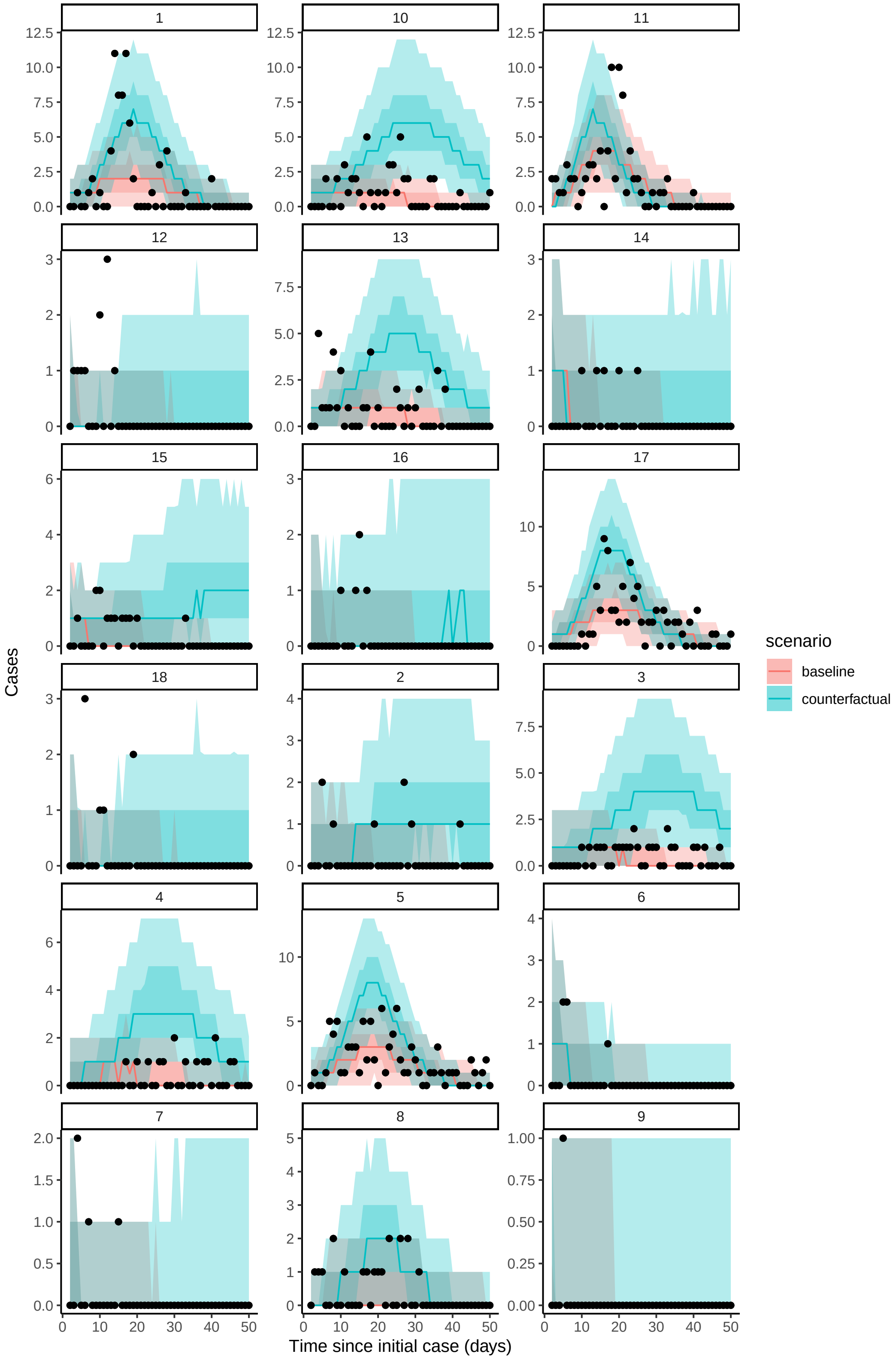


Figure S3: Counterfactual scenarios from hierarchical $R_{0,k}$ model where there is no intervention (baseline, $\zeta = 0$) compared to model with intervention.

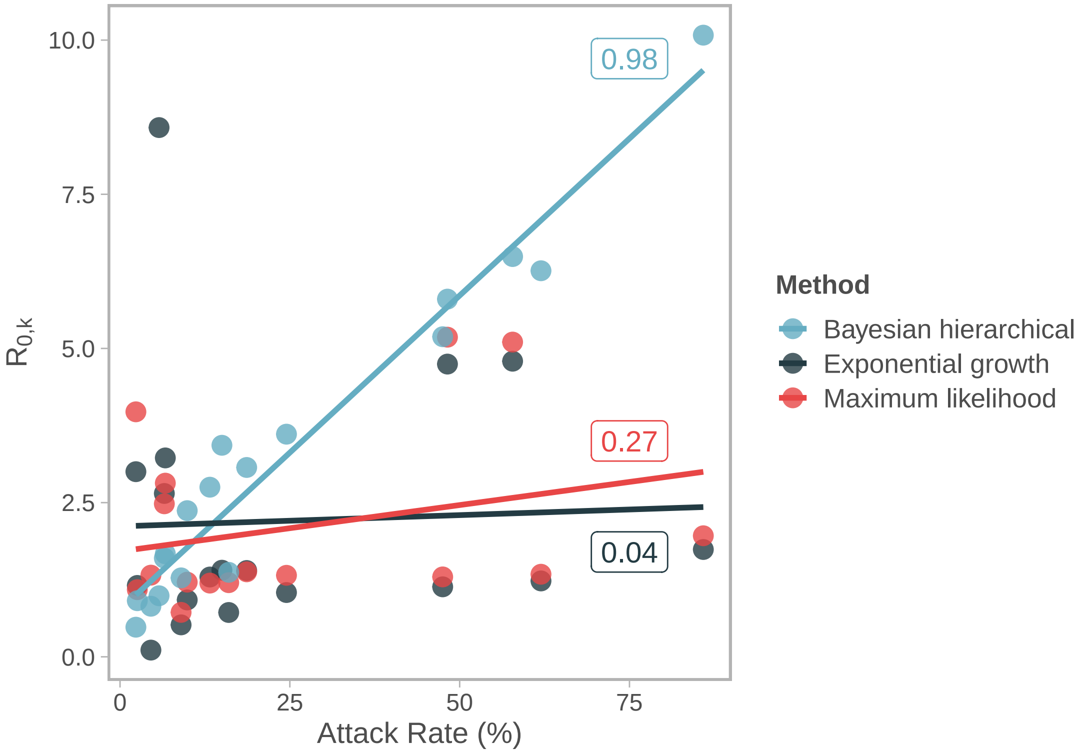


Figure S4: **Scatterplot of $R_{0,k}$ estimates against attack rate, with Pearson correlation coefficients and linear regression.**

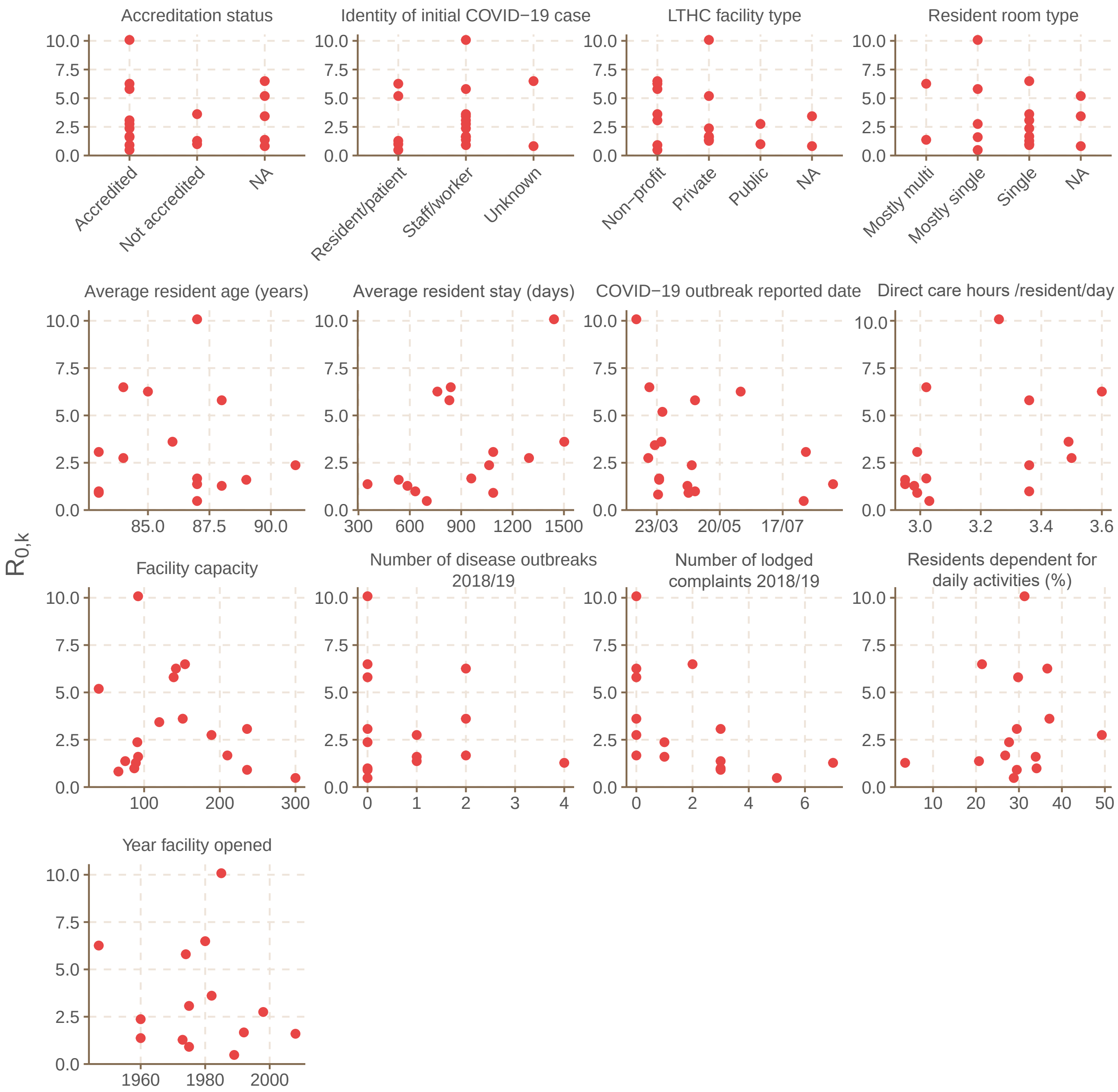


Figure S5: **Relationship between $R_{0,k}$ estimates and various LTHC facility factors.** Non-COVID-19 data is from the 2018/19 year [S1].

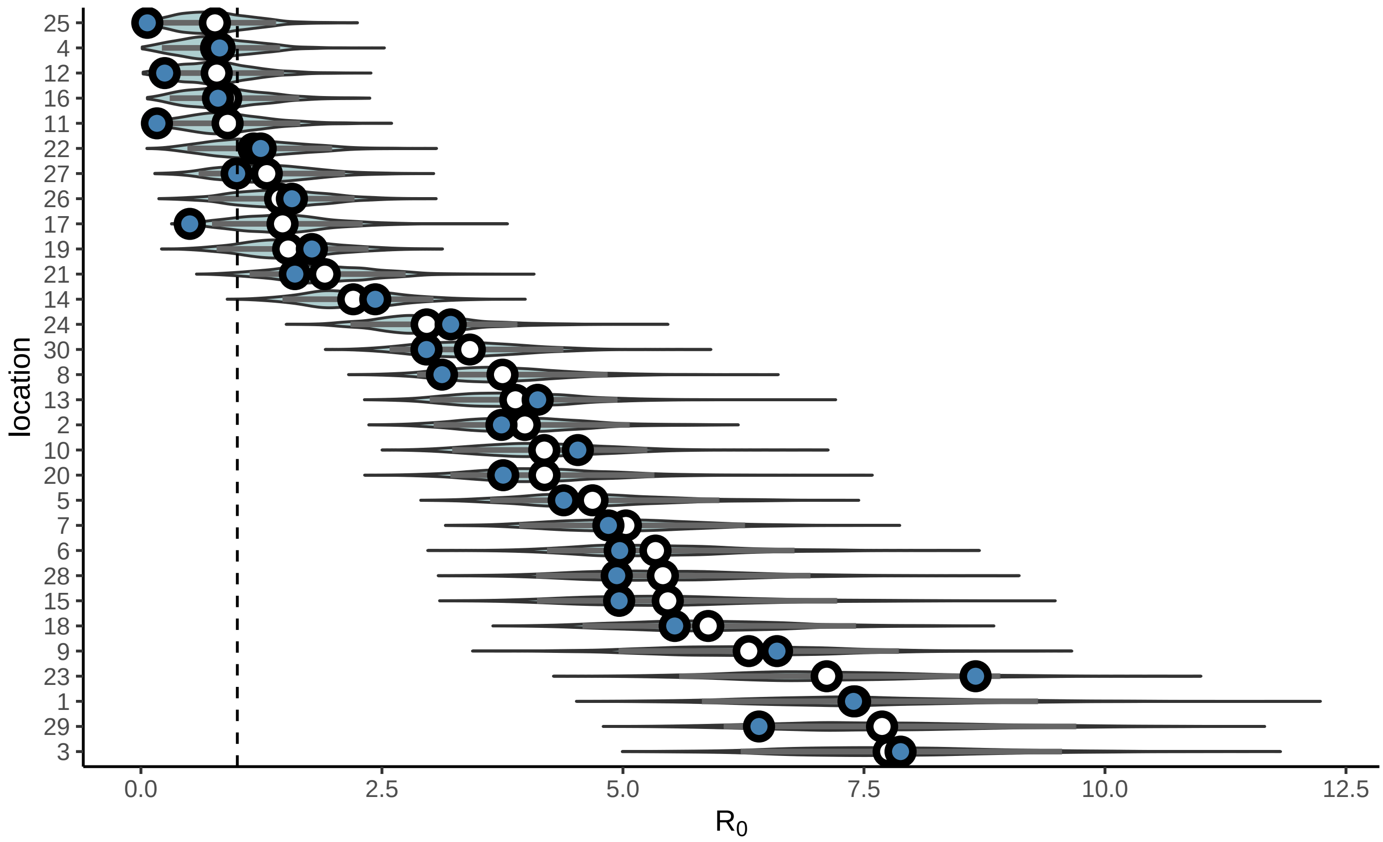


Figure S6: **Validation of hierarchical Bayesian procedure on simulated data.** $R_{0,k}$ values for each simulated outbreak were drawn from a truncated normal distribution with $\mu = 3, \sigma = 3$, with truncation at 0. An SEIR model with fixed intervention was performed with Poisson drawn incidence data. Estimates are shown as white points with shaded 95% credible intervals with true values shown as blue points.

### References

[S1] Office of the Seniors Advocate, British Columbia . Long-Term Care Facilities Quick Facts Directory. https://www.seniorsadvocatebc.ca/quickfacts/location; 2020. Accessed: November 30th, 2020.
